# COVID-19 hospitalizations in Brazil’s Unified Health System (SUS)

**DOI:** 10.1101/2020.09.03.20187617

**Authors:** Carla Lourenço Tavares de Andrade, Claudia Cristina de Aguiar Pereira, Mônica Martins, Sheyla Maria Lemos Lima, Margareth Crisóstomo Portela

**Affiliations:** Escola Nacional de Saúde Pública Sergio Arouca, Fundação Oswaldo Cruz

**Keywords:** COVID-19, Hospitalizations, Hospital mortality, SUS

## Abstract

**Objective:** To study the profile of hospitalizations due to COVID-19 in the Unified Health System (SUS) in Brazil and to identify factors associated with hospital mortality related to the disease.

**Methods:** Cross-sectional study, based on secondary data on COVID-19 hospitalizations that occurred in SUS, between the last days of February and June. Patients aged 18 years or older, with primary or secondary diagnoses indicative of COVID-19 were included. Bivariate analyses were performed and generalized linear mixed models (GLMM) were estimated with random effects intercept. The modeling followed three steps, including: attributes of the patients; elements of the care process; and characteristics of the hospital and place of hospitalization.

**Results:** 89,405 hospitalizations were observed, of which 24.4% resulted in death. COVID-19 patients hospitalized in SUS were predominantly male (56.5%), with a mean age of 58.9 years. The length of stay ranged from less than 24 hours to 114 days, with a mean of 6.9 (±6.5) days. Of the total number of hospitalizations, 22.6% reported ICU use. The chances of hospital death among men were 16.8% higher than among women and increased with age. Black individuals had a higher chance of death. The behavior of the Charlson and Elixhauser indices was consistent with the hypothesis of a higher risk of death among patients with comorbidities, and obesity had an independent effect on increasing this risk. Some states had a higher risk of hospital death from COVID-19, such as Amazonas and Rio de Janeiro. The chances of hospital death were 72.1% higher in municipalities with at least 100,000 inhabitants and being hospitalized in the municipality of residence was a protective factor.

**Conclusion:** There was wide variation in hospital COVID-19 mortality in the SUS, associated with demographic and clinical factors, social inequality and differences in the structure of services and quality of health care.

## Introdução

The Covid-19 pandemic caused by the SARS-CoV-2 virus has severely affected Brazil, which has become the country with the second highest number of cases and deaths in the world^1^. The first confirmed case of COVID-19 in Brazil and Latin America occurred on February 26, 2020 in the state of São Paulo. Less than a month later, the first death also occurred in São Paulo on March 17. Social distancing measures were first introduced in March in five states, chronologically as follows: Goiás, Rio de Janeiro, Santa Catarina, Distrito Federal and São Paulo^2^. Due to the rapid spread of the disease, all 26 states and the Federal District had already registered ten or more cases of the disease in early April, with a higher concentration of cases in the Southeast Region, especially in the states of São Paulo and Rio de Janeiro^3^.

Countries with universal health systems and which implemented effective measures of social distancing early on seem to have had better results in terms of number of cases and deaths. Given the rapid transmission of the virus, there was an abrupt and growing additional demand for hospitalizations worldwide, thus putting health care systems under strain in many countries^4^. According to the World Health Organization (WHO), 80% of patients with COVID-19 have mild and uncomplicated symptoms, 15% progress to hospitalization and 5% require admission to the intensive care unit (ICU)^5^.

Brazil’s Unified Health System (SUS) is the largest public and universal health system in the world, which penetrates the entire country, covering approximately 75% of the population that does not have private health insurance^6,7^. The system’s underfunding and inadequate management, however, has undermined the ability to offer health services, with wide variations in the quality of services provided. The need to cope with COVID-19 has shown weaknesses in the system, despite the increase in the number of general and intensive care hospital beds offered and the construction of field hospitals. Several Brazilian states had to deal, to a greater or lesser extent, with a demand greater than the response capacity of SUS, with long queues for general beds and intensive care.

In addition to the structure of the services supplied for COVID-19 cases and the measures implemented to control the pandemic, patient characteristics, including age, sex, race/color and pre-existing conditions, interfere with the demand for hospitalization, the care provided and the outcomes of these hospitalizations^8^.

The literature about COVID-19 hospital mortality has been consistent around the world. It is known from a systematic review and meta-analysis that, in hospitalizations of patients aged 18 years and older, ICU mortality due to COVID-19 is higher than that normally seen in patients with other viral pneumonias. As the pandemic progressed, reported death rates dropped from more than 50% to close to 40%. There were no significant effects by geographic location. Countries that faced the pandemic later appear to have better dealt with cases, but in order to better understand the mechanisms, it would be necessary to study the admission criteria applied, patient status upon admission and treatments administered in the ICU^4^. Another systematic review carried out on inpatients diagnosed with COVID-19 in 12 countries showed that patients in America and Europe were older than those in Asia. In Europe there were more male patients. Comorbidities were more frequent, and mortality was higher in European and American patients than in Asian patients. Despite considerable variations in the reviewed studies due to sample size, disease severity (mild vs. critical illness) and geographic region, the most frequent comorbidities were obesity, hypertension, diabetes mellitus, cardiovascular, lung, chronic kidney and liver diseases, immunosuppression and cancers.

In addition, socioeconomic status of the affected population also plays a role in these results. For instance, a study carried out in New York, using a cohort of patients hospitalized with COVID-19 coming from a poor population, identified severe obesity as a risk factor for higher mortality, even after adjusting for other confounding factors^9^.

In Brazil, a cross-sectional observational study conducted with hospitalized COVID-19 patients identified a lower likelihood of death in young, female patients with fewer comorbidities, commensurable with what has been observed in other countries. Furthermore, it highlighted a higher risk of death among black and brown populations and in patients hospitalized in the Northern region compared to other regions of the country, which, for the authors, would be a specific manifestation of the disease in the Brazilian population. The authors suggest that the regional effect may be driven by the morbidity profile of patients in regions with lower levels of socioeconomic development^10^.

Despite the recognized increase in international and national publications about COVID-19, there are still few studies that explore the risk factors and characteristics of hospitalizations for COVID-19 in the population considering geographic variations^11^. Therefore, the following question arises: How does the profile of patients and hospitalizations observed in the international literature compare with hospitalizations that occurred in Brazil’s public system in different regions of the country? The goal of this paper is to understand the profile of COVID-19 hospital admissions in the Unified Health System (SUS) and to identify associated factors with the occurrence of hospital deaths related to the disease, considering patient characteristics and the care offered, with a focus on regional variations. In this way, we expect to contribute to the improvement of the care provided and to define strategies to face future developments in the pandemic’s course until the population has access to an effective vaccine.

## Methods

### Study Design

This is a cross-sectional, observational study based on secondary data on COVID-19 hospitalizations that occurred in the Unified Health System (SUS), which were available on the DATASUS website on August 4, 2020^12^, considering the first four months of the pandemic in Brazil – between the end of February and the last week of June.

The SUS Hospital Information System (SIH) was the data source. Although this system presents coverage and quality issues, it is the main source of information on hospital production nationwide. In addition to demographic data (age, sex), it includes diagnostic data, type of admission (elective / emergency) and type of care (surgical / clinical), length of stay (LOS), use of intensive care (ICU), outcome at discharge and the amount reimbursed for the hospitalization. In 2016, it was expanded to accept up to nine secondary diagnoses registrations, potentially allowing a better description of the morbidity and severity case profile. In addition to secondary diagnoses, variables indicating their pre-existence (presence at admission), which are patient attributes; or if they were acquired during the process of care in the hospital, which recognizably are performance and quality of care issues. The only sociodemographic variable available in the SIH dataset is race/color.

The data used were extracted from the reduced type (RD) files for each state and Federal District, freely available on the DATASUS portal^12^. It is likely that hospitalizations in the months of interest are underreported due to the specificities of SIH’s data transmission process and its main purpose, which is reimbursement.

### Study sample

Initially, we excluded hospitalizations of patients under 18 years of age. The selection of COVID-19 hospitalizations began with the following variables: procedure performed (which designates the type of treatment performed on the patient, whether clinical or surgical and serves as the basis for hospitalization payment), primary and secondary diagnoses. We considered as cases those patients whose hospital record indicated an association with COVID-19 in any of these variables. Thus, all hospitalizations with the primary diagnosis or one of the secondary diagnoses identified with the International Statistical Classification of Diseases and Related Health Problems, 10th revision (ICD-10) code B34.2 – coronavirus infection of unspecified location – were included. This diagnostic category was defined in the technical guidelines of the SIH in the context of the pandemic^13^. Also following these guidelines, we included hospitalizations coded as “03.03.01.022–3 - TREATMENT OF INFECTION BY THE NEW CORONAVIRUS - COVID 19” in the procedure variable. This code was recently created to take effect from April 14, 2020 onwards. In hospitalizations prior to April, the procedure used was “03.03.01.019-3 - TREATMENT OF OTHER DISEASES CAUSED BY VIRUSES (ICD-10: B25-B34)”. Given its lack of specificity and relation to a wide range of main diagnoses, it was not considered as an additional inclusion criterion beyond the B34.2, except for a few records containing the B97.2 - Coronavirus, as the cause of classified diseases in other chapters as the main diagnosis.

### Data analyses

The study focused on the analysis of patient sociodemographic and clinical characteristics, the care process and contextual variables related to the hospitalization, and their effects on the chances of hospital death.

Descriptive and bivariate analyses were performed to characterize the study population and to test the relationships between the independent variables (attributes of the patients, the care process and the hospital, place of residence and geographic location) with the dependent variable ‘in-hospital death’. The scope of the information available in the dataset shaped the range of operationalized variables and the scope of the analyses.

To account for the correlation among observations occurring in each hospital, resulting from the process of care and case-mix, we used the generalized linear mixed model (GLMM) with random effects intercept. This model was used to assess the different factors associated (independent variables) with the death of hospitalized COVID-19 patients (response with binary distribution). Thus, the modeling occurred in three stages, with the insertion of different blocks of variables: (i) patient attributes - variables that express the severity of the case or worse prognosis, and social inequality (race / color); (ii) elements of the care process; and (iii) characteristics of the hospital, place of residence and place of hospitalization.

For the first stage, the case severity profile was based on demographic variables (sex and age) and comorbidities. The variable ‘sex’ is binary as informed in the SIH, and we considered female as the reference category. Age was treated as a categorical variable (18-39, 40-49, 50-59, 60-69, 70-79, 80-89 and ≥ 90 years). Comorbidities were contemplated in different ways: (i) calculating the Charlson Comorbidity Index (CCI)^15^; (ii) identifying presence of the comorbidities as proposed by Elixhauser Comorbidity Index (ECI) for administrative databases^16^; (ii) considering specific comorbidities - obesity, arterial hypertension and diabetes^17^ due to their frequency in the population and their relevance in the COVID-19 literature, although they are already a part of the previous comorbidity measures used. CCI and ECI were chosen because they are measures widely used in models for predicting death^17^ and have previously been applied to Brazilian patients^18^; the calculation used the ICD-10 code algorithm for each clinical condition developed by Quan et al.^19^. Additionally, we considered whether COVID-19 was designated as a primary (reference category) or secondary diagnosis. In the case of the variable ‘race/color’, we started out by using the five categories provided by SIH (white, black, brown, yellow and indigenous) and, after examination, we chose to consider the categories black, brown and others, as reference. Race/color is used as a proxy for social vulnerability and inequalities in socioeconomic conditions, health, access, use and effectiveness of care.

In the second stage, two variables about the care process were added to the previous model: ICU use and LOS. The number of days spent in the intensive care unit (ICU) was transformed into a dichotomous variable (yes / no). The LOS was also categorized considering the distribution of deaths. Therefore, the following categories were built: 1 day or less; 2-7 days; 8-22 days and ≥ 23 days; hospitalizations whose LOS was 0, were considered to be less than 24 hours and included in the first category.

In the third and last stage of the modeling process, we inserted hierarchical level variables, namely the hospital, the place of residence and the geographical location where the hospitalization occurred. At the hospital level, its legal nature was also considered, and they were categorized as municipal, state and federal public hospital, private for profit and private non-profit. For the place of residence, a variable indicating patient displacement to seek care was created, considering whether the municipality of residence and the location of the hospital were the same (dichotomous variable yes no). Finally, we used categories for each state (including the Federal District) to account for geographic effects in the occurrence of hospitalization; and municipal population size, which, in previous analysis, was more adequate than the municipal human development index (Municipal HDI).

The goodness of fit of the different models was tested. Predictive capacity was assessed based on the “c” statistic. Data analysis was performed with the SAS statistical package.

## Results

The selection criteria for hospitalizations yielded 89,405 records, among which 13 corresponded to hospitalizations that started and ended in 2020, before the month of March. Altogether, 21,807 (24.4%) hospitalizations resulted in death.

The COVID-19 patients hospitalized in SUS were predominantly male (56.5%), aged between 18 and 114 years old, mean and standard deviation of 58.9 (± 16.8) and median of 60 years. The LOS ranged from 24 hours or less to 114 days, with an average of 6.9 (± 6.5) days and a median of 5 days. Altogether, hospitalizations represented R$ 332.10 million (Brazilian reais) in expenditures for SUS, which varied between 40.40 and 111,914.50 reais per patient, with an average of R$ 3,714.50 (± 6,119.20) and first, second and third quartiles were, 1,500.00, 1,610.40 and 2,087.20 Brazilian reais, respectively.

From the total number of hospitalizations, 22.6% registered ICU use, which accounted for 66.4% of the total amount paid to all COVID-19 hospitalizations. In these admissions, the average and median hospital stay time was 10.3 (± 8.5) days and 8 days, respectively, and the amount paid for average and median hospital stay was respectively 11,083.10 (± 9,674.30) and 8,036.40 reais. Considering ICU alone, the average LOS was 7.6 (± 6.8) days, with a median of 6 days.

The analyses here presented considered the association between the occurrence of hospital death and three “blocks” of variables: sociodemographic and clinical attributes of patients; aspects related to the care process; and data from the macro context of the hospital organization and geographic location of the hospital. Tables 1-3 show descriptive statistics and bivariate analyses of these variables, when death occurred or not.

**Table 1.** Bivariate analyses of sociodemographic and clinical variables and hospital mortality. COVID-19 hospitalizations in the Unified Health System in Brazil (N = 89,405). February to June 2020

| Variables | N | % | Hospital death | | | | $\chi^2$<br>(p-value) |
| --- | --- | --- | --- | --- | --- | --- | --- |
|  |  |  | Yes |  | No |  |  |
|  |  |  | N | % | N | % |  |
| Sex |  |  |  |  |  |  | < 0.0001 |
| Male | 50,520 | 56.5 | 12,867 | 25.5 | 37,653 | 74.5 |  |
| Female | 38,885 | 43.5 | 8,940 | 23.0 | 29,945 | 77.0 |  |
| Age (years) |  |  |  |  |  |  | < 0.0001 |
| 18-39 | 13,028 | 14.6 | 1,113 | 8.5 | 11,915 | 91.5 |  |
| 40-49 | 13,556 | 15.2 | 1,730 | 12.8 | 11,826 | 87.2 |  |
| 50-59 | 17,724 | 19.8 | 3,234 | 18.2 | 14,490 | 81.8 |  |
| 60-69 | 19,098 | 21.4 | 5,363 | 28.1 | 13,735 | 71.9 |  |
| 70-79 | 15,593 | 17.4 | 5,615 | 36.0 | 9,978 | 64.0 |  |
| 80-89 | 8,483 | 9.5 | 3,775 | 44.5 | 4,708 | 55.5 |  |
| ≥ 90 | 1,923 | 2.1 | 977 | 50.8 | 946 | 49.2 |  |
| Ethnic group |  |  |  |  |  |  | < 0.0001 |
| White | 21,260 | 23.8 | 4,635 | 21.8 | 16,625 | 78.2 |  |
| Black | 5,507 | 6.2 | 1,754 | 31.9 | 3,753 | 68.1 |  |
| Pardo | 33,542 | 37.5 | 8,741 | 26.1 | 24,801 | 73.9 |  |
| East Asian | 3,071 | 3.4 | 595 | 19.4 | 2,476 | 80.6 |  |
| Indigenous | 152 | 0.2 | 44 | 28.9 | 108 | 71.1 |  |
| Ignored | 25,873 | 28.9 | 6,038 | 23.3 | 19,835 | 76.7 |  |
| Charlson Index |  |  |  |  |  |  | < 0.0001 |
| 0 | 86,131 | 96.3 | 20,560 | 23.9 | 65,571 | 76.1 |  |
| 1 | 2,552 | 2.9 | 898 | 35.2 | 1,654 | 64.8 |  |
| ≥ 2 | 722 | 0.8 | 349 | 48.3 | 373 | 51.7 |  |
| Elixhauser comorbidities |  |  |  |  |  |  | < 0.0001 |
| Yes | 5,574 | 6.2 | 1,887 | 33.9 | 3,687 | 66.1 |  |
| No | 83,831 | 93.8 | 19,920 | 23.8 | 63,911 | 76.2 |  |
| Obesity |  |  |  |  |  |  | < 0.0001 |
| Yes | 655 | 0.7 | 224 | 34.2 | 431 | 65.8 |  |
| No | 88,750 | 99.3 | 21,583 | 24.3 | 67,167 | 75.7 |  |
| Hypertension |  |  |  |  |  |  | < 0.0001 |
| Yes | 3,794 | 4.2 | 1,273 | 33.6 | 2,521 | 66.4 |  |
| No | 85,611 | 95.8 | 20,534 | 24.0 | 65,077 | 76.0 |  |
| Diabetes |  |  |  |  |  |  | < 0.0001 |
| Yes | 1,302 | 1.5 | 424 | 32.6 | 878 | 67.4 |  |
| No | 88,103 | 98.5 | 21,383 | 24.3 | 66,720 | 75.7 |  |
| COVID-19 as secondary diagnosis |  |  |  |  |  |  | 0.9408 |
| Yes | 2,777 | 3.1 | 679 | 24.5 | 2,098 | 75.5 |  |
| No | 86,628 | 96.9 | 21,128 | 24.4 | 65,500 | 75.6 |  |

Source: Ministry of Health – SUS' Inpatient Care Information System

**Table 2.**
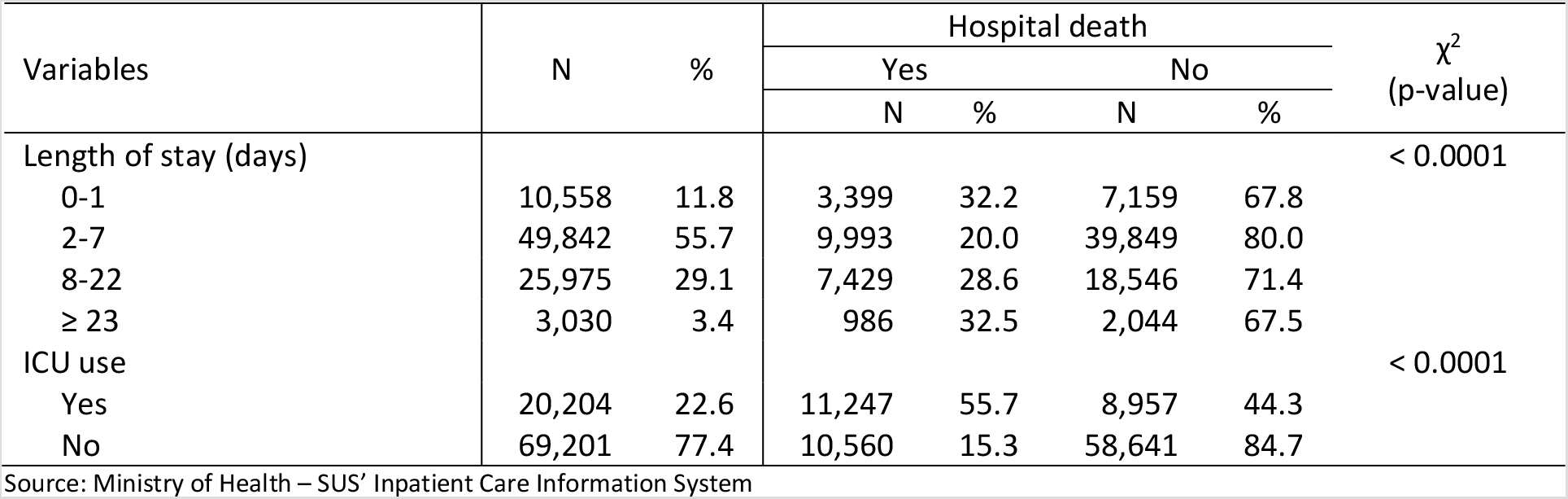
Bivariate analyses of variables related to inpatient healthcare process and hospital mortality. COVID-19 hospitalizations in the Unified Health System in Brazil (N = 89,405). February to June 2020

**Table 3.** Bivariate analyses of macro context variables and hospital mortality. COVID-19 hospitalizations in the Unified Health System in Brazil (N = 89,405). February to June 2020

| Variables | N | % | Hospital death | | | | $\chi^2$<br>(Valor de p) |
| --- | --- | --- | --- | --- | --- | --- | --- |
|  |  |  | Yes |  | No |  |  |
|  |  |  | N | % | N | % |  |
| Hospital ownership |  |  |  |  |  |  | < 0.0001 |
| Public – Municipality | 37,539 | 42.0 | 7,544 | 20.1 | 29,995 | 79.9 |  |
| Pubic – State | 28,728 | 32.1 | 8,903 | 31.0 | 19,825 | 69.0 |  |
| Public – Federal | 958 | 1.1 | 313 | 32.7 | 645 | 67.3 |  |
| Private | 4,119 | 4.6 | 862 | 20.9 | 3,257 | 79.1 |  |
| Philanthropic | 18,061 | 20.2 | 4,185 | 23.2 | 13,876 | 76.8 |  |
| State |  |  |  |  |  |  | < 0.0001 |
| Rondônia | 957 | 1.1 | 127 | 13.3 | 830 | 86.7 |  |
| Acre | 64 | 0.1 | 28 | 43.8 | 36 | 56.3 |  |
| Amazonas | 4,682 | 5.2 | 1,628 | 34.8 | 3,054 | 65.2 |  |
| Roraima | 677 | 0.8 | 243 | 35.9 | 434 | 64.1 |  |
| Pará | 3,398 | 3.8 | 647 | 19.0 | 2,751 | 81.0 |  |
| Amapá | 237 | 0.3 | 106 | 44.7 | 131 | 55.3 |  |
| Tocantins | 265 | 0.3 | 55 | 20.8 | 210 | 79.2 |  |
| Maranhão | 5,946 | 6.6 | 1,644 | 27.6 | 4,302 | 72.4 |  |
| Piauí | 1,060 | 1.2 | 115 | 10.8 | 945 | 89.2 |  |
| Ceará | 6,141 | 6.9 | 1,472 | 24.0 | 4,669 | 76.0 |  |
| Rio Grande do Norte | 917 | 1.0 | 238 | 26.0 | 679 | 74.0 |  |
| Paraíba | 848 | 0.9 | 166 | 19.6 | 682 | 80.4 |  |
| Pernambuco | 8,524 | 9.5 | 2,370 | 27.8 | 6,154 | 72.2 |  |
| Alagoas | 804 | 0.9 | 246 | 30.6 | 558 | 69.4 |  |
| Sergipe | 271 | 0.3 | 44 | 16.2 | 227 | 83.8 |  |
| Bahia | 2,853 | 3.2 | 780 | 27.3 | 2,073 | 72.7 |  |
| Minas Gerais | 2,987 | 3.3 | 500 | 16.7 | 2,487 | 83.3 |  |
| Espírito Santo | 1,932 | 2.2 | 559 | 28.9 | 1,373 | 71.1 |  |
| Rio de Janeiro | 10,589 | 11.8 | 3,401 | 32.1 | 7,188 | 67.9 |  |
| São Paulo | 28,396 | 31.8 | 6,235 | 22.0 | 22,161 | 78.0 |  |
| Paraná | 1,753 | 2.0 | 291 | 16.6 | 1,462 | 83.4 |  |
| Santa Catarina | 988 | 1.1 | 147 | 14.9 | 841 | 85.1 |  |
| Rio Grande do Sul | 2,173 | 2.4 | 325 | 15.0 | 1,848 | 85.0 |  |
| Mato Grosso do Sul | 81 | 0.1 | 7 | 8.6 | 74 | 91.4 |  |
| Mato Grosso | 384 | 0.4 | 76 | 19.8 | 308 | 80.2 |  |
| Goiás | 902 | 1.0 | 124 | 13.7 | 778 | 86.3 |  |
| Distrito Federal | 1,576 | 1.8 | 233 | 14.8 | 1,343 | 85.2 |  |
| Population size |  |  |  |  |  |  | < 0.0001 |
| ≤ 50.000 hab. | 8,671 | 9.7 | 878 | 10.1 | 7,793 | 89.9 |  |
| 50.001-100.000 hab. | 7,639 | 8.5 | 1,573 | 20.6 | 6,066 | 79.4 |  |
| 100.001-500.000 hab. | 23,442 | 26.2 | 5,785 | 24.7 | 17,657 | 75.3 |  |
| > 500.000 hab. | 49,653 | 55.5 | 13,571 | 27.3 | 36,082 | 72.7 |  |
| Residence/hospitalization |  |  |  |  |  |  | < 0.0001 |
| Same municipality | 68,069 | 76.1 | 15,565 | 22.9 | 52,504 | 77.1 |  |
| Different municipalities | 21,336 | 23.9 | 6,242 | 29.3 | 15,094 | 70.7 |  |

Source: Ministry of Health – SUS' Inpatient Care Information System

In Table 1, we observe higher hospital mortality for men and an increasing gradient of the odds of dying as age (age group) increases. Among individuals aged 18 to 39 years, 8.5% of hospitalizations resulted in death. For individuals between 70 and 79, 80 and 89 and at least 90 years old, the proportion of hospitalizations that resulted in death increased to 36.0%, 44.5% and 50.8%, respectively.

Brown individuals are the majority in the ‘race/color’ variable, but the high percentage of hospitalizations with missing ‘race/color’ data (28.9%) draws special attention. Among the cases with non-missing information on the variable, there is a higher occurrence of deaths among blacks (31.9%), followed by indigenous people (28.9%) and browns (26.1%). The percentage of hospital deaths with the ‘race/color’ missing data is slightly higher than that observed among whites and lower than that observed among browns. It is also worth noting that only 152 admissions of indigenous individuals were observed, making up 0.2% of the total.

The clinical variables expose low levels of information about secondary diagnoses, resulting in a significant underreporting of clinical conditions relevant to the patients’ prognosis. Approximately 78.3% of hospitalizations did not have any secondary diagnoses. Even so, the results indicate a higher occurrence of deaths among patients with greater severity according to the Charlson and Elixhauser comorbidity indices or patients with diseases such as hypertension, diabetes and obesity, which show a pattern consistent with what was expected.

Table 1 also shows a variable created to capture any differences in mortality between patients with COVID-19 as the main diagnosis or as a secondary diagnosis. Hospitalizations with the COVID-19 diagnosis registered in one of the secondary diagnosis corresponded to 3.1% of the sample, and in this case, the bivariate analyses did not show differences regarding the occurrence of death.

In Table 2, the percentage of hospitalizations with less than 24 hours or one day of hospitalization stands out (11.8%), and moreover, the high mortality (32.2%) in this category. About two thirds of the total hospitalizations lasted up to 7 days (29.1%), between 8 and 22 days (3.4%) and 23 days or more (3.4%). Based on the bivariate analyses, there appears to be a greater concentration of deaths in the extreme categories of LOS, with a lower occurrence of deaths (20.0%) among those with LOS between 2 and 7 days. The proportion of deaths among those who used the ICU was high (55.7%) compared to those who did not use it (15.3%).

It is worth highlighting the difference between the LOS between those who did not and did die during hospitalization. In the first group, the average LOS was 6.7 (± 6.2) days, and the median was 5 days. Among the patients who died, the average LOS was 7.6 (± 7.2) days, and the median was 6 days.

Table 3 shows that most hospitalizations for COVID-19 occurred in municipal public hospitals (42%), followed by state public hospitals (32.1%) and philanthropic hospitals (20.2%). The share of private hospitals contracted by SUS was 4.6%, while that of federal hospitals was 1.1%.

The distribution of hospitalizations by state presents, in a way, a photograph of the period analyzed, in which some states in the Southeastern, Northeastern and Northern regions were most affected by the epidemic. Just under a third (31.8%) of the hospitalizations analyzed occurred in São Paulo, the largest and richest Brazilian state. São Paulo was followed by Rio de Janeiro (11.8%), Pernambuco (9.5%), Ceará (6.9%), Maranhão (6.7%) and Amazonas (5.2%). Among these states, the occurrence of hospital deaths was especially high in Amazonas (34.1%) and Rio de Janeiro (32.1%), with figures also higher than the national average (24.4%) in Pernambuco (27.8%) and Maranhão (27.6%). The Amazonian states of Acre, Roraima and Amapá corresponded, respectively, to 0.1%, 0.8% and 0.3% of hospitalizations in the country, but they are states with relatively small populations, which stood out for the high proportions of observed hospital deaths - 43.8%, 35.9% and 44.7%, respectively. The states of Alagoas (30.6%), Espírito Santo (28.9%), Bahia (27.3%) and Rio Grande do Norte (26, 8%) also had hospital death percentages higher than the national average.

Table 3 also shows that 81.7% of admissions for COVID-19 in the first four months of the epidemic in the country occurred in municipalities with more than 100 thousand inhabitants, with more deaths observed in these municipalities than in other smaller ones. More than ¾ of hospitalizations were carried out in hospitals located in the same municipality of residence of the patient, with a higher occurrence of deaths when the patient had to travel to receive hospital care.

Table 4 presents the three regression models that explain the occurrence of hospital death, considering the progressive inclusion of the “blocks” of variables already mentioned. The first line of the table provides the variance of random intercepts related to hospital units for each model.

**Table 4.**
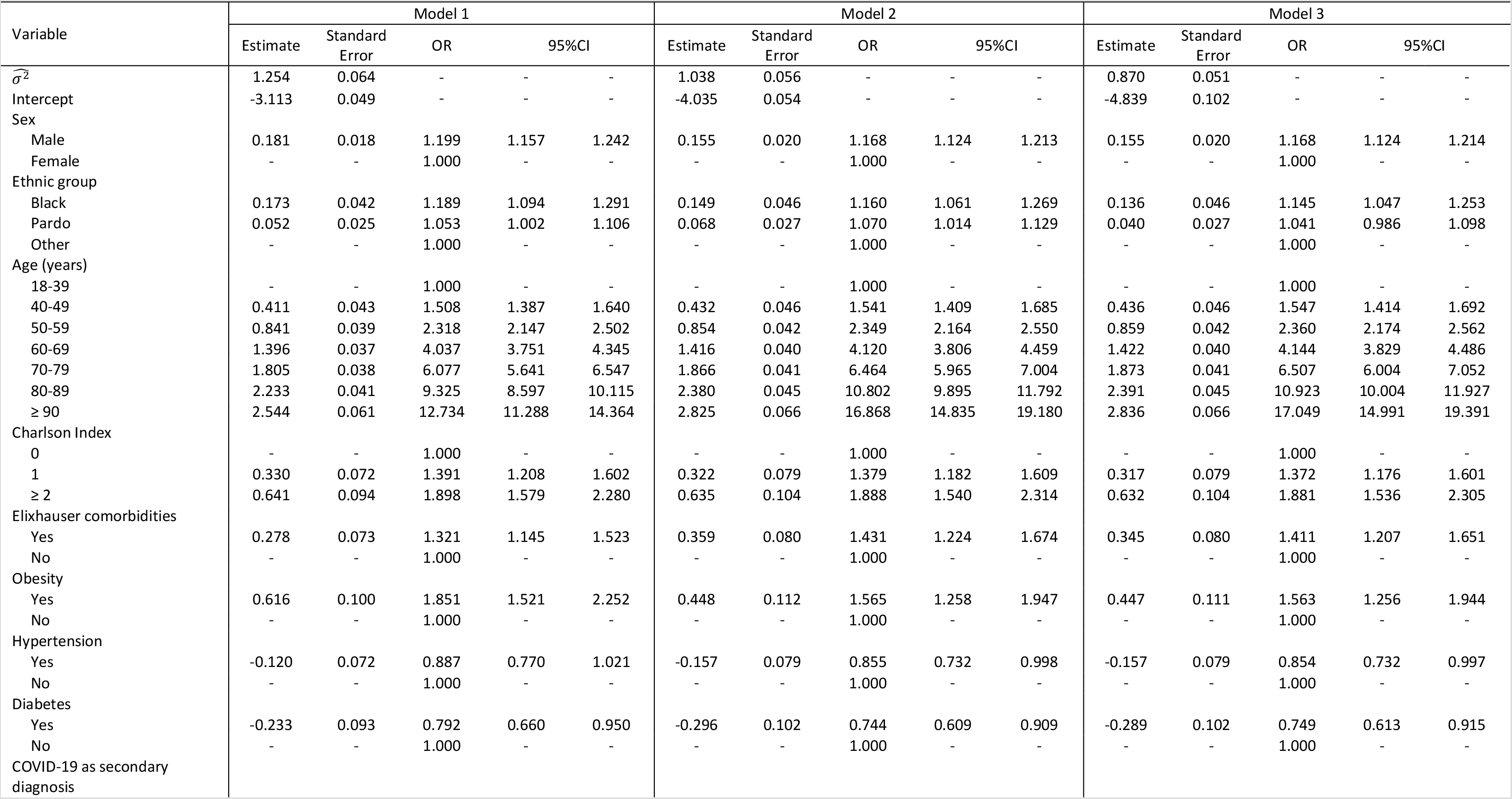
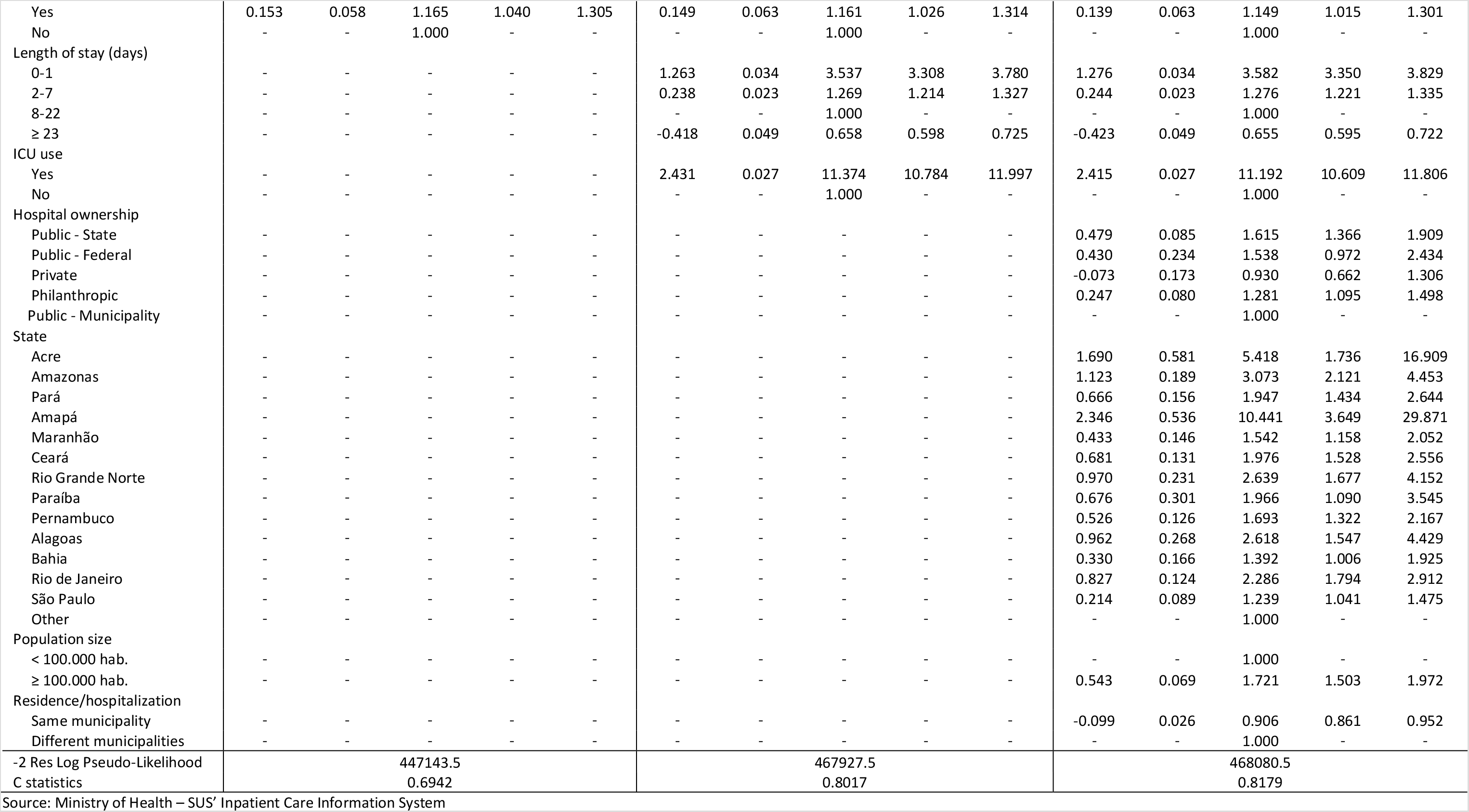
Multivel logistic regression models explaining the variation in hospital mortality in COVID-19 hospitalizations in the Unified Health System (N = 89,405). Brazil, February to June 2020

In general terms, we observe that some patterns of mortality among categories of variables change from the descriptive analyses to the multivariate models, given the control for confounding factors. There is also consistency in the results from adding “blocks” of variables from one model to the next, although, strictly speaking, the variable ‘race/color’ brown loses statistical significance between the second and third models.

Considering model 3, it is possible to observe that the odds of hospital death among men were 16.8% higher than among women. The patient’s age group was also an important predictor of the likelihood of death. Compared to patients between 18 and 39 years old, patients aged 40 to 49 years were 54.7% more likely to die in the hospital, while for patients aged 90 years or more this increase reached 1,604.9%. Furthermore, blacks had higher odds of death during the hospital stay (OR = 1.14; 95% CI 1.05-1.25), compared to the reference category including whites, yellows, indigenous and individuals without a record of the variable. The adjusted risk of hospital death for browns was not statistically significant at the 5% level, but “borderline”.

Despite the substantial underreporting of clinical conditions, the behavior of the Charlson and Elixhauser indices was consistent with the hypothesis of higher odds of death among patients with comorbidities. The odds of death were 37.2% and 88.1% (model 3) higher among patients with Charlson index scores equal to 1 and ≥2, respectively, compared to those with score equal to zero, controlling for other variables. For those with Elixhauser comorbidities, the adjusted odds of dying were 41.1% higher than for those without such comorbidities. Presence of hypertension (OR = 0.85; 95% CI 0.73-0.99) and diabetes (OR = 0.75; 95% CI 0.61-0.91) had protective effects, regardless of the inclusion of the other comorbidity measures. Obesity was statistically associated with higher odds of hospital death (56.3% higher among obese people, compared to non-obese people). Finally, patients who had COVID-19 as a secondary diagnosis, liable, in some cases, to have acquired the infection in the hospital itself, had 14.9% higher odds of dying in the hospital than those who had the disease registered as the main diagnosis.

Some changes were observed in the second regression model when compared to what was observed in the bivariate analyses of LOS and occurrence of death (Table 2). Controlling for other variables, the higher odds of hospital death (OR = 3.58; 95% CI 3.35-3.83) for those whose stay was up to 1 day remained evident. Compared to patients with LOS between 8 and 22 days, those with LOS between 2 and 7 days were more likely to die (OR = 1.28; 95% CI 1.22-1.34), while patients with a hospital stay of at least 23 days were less likely (OR = 0.66; 95% CI 0.60-0.72). ICU use was a relevant predictor of higher odds of hospital death (OR = 11.19; 95% CI 10.61-11.81).

Regarding the third model, which included contextual variables related to the organizational aspect of the hospital and its geographic location, the results point to greater odds of hospital deaths in state public hospitals and philanthropic hospitals, compared to municipal public hospitals, in addition to a group of Brazilian states where hospital mortality due to COVID-19 was more critical in the period studied. The hospitalized patients in the states of Amazonas, Rio Grande do Norte, Alagoas, Rio de Janeiro, Ceará, Paraíba, Pará, Pernambuco and Maranhão, were at least 50% more likely to die than in other states in the reference category (predominantly states in the South and Midwest regions), controlling for other variables. The highest odds ratios were, however, observed for Acre (OR = 5.42; 95% CI 1.74-16.91) and Amapá (OR = 10.44; 95% CI 3.65-29.87), which, due to the small number of hospitalizations, had estimates with very wide confidence intervals.

Finally, the odds of death during hospitalization were 72.1% higher in municipalities with at least 100 thousand inhabitants and being admitted to a hospital in the same municipality of residence remained a protective factor for the outcome variable considered.

Based on the statistics related to the goodness of fit of the models, the three blocks of variables in the models constituted explanatory factors for the variation in hospital deaths. The attributes of the patients themselves allowed for a relatively high predictive capacity of the model (c = 0.69), which significantly increased by the inclusion of variables related to the care process (c = 0.80) and then had a modest increase when including variables related to the hospital’s organizational context and geographic area (c = 0.82).

## Discussion

The study provides a comprehensive overview of COVID-19 hospitalizations that occurred in the SUS, including 89,405 hospitalizations, among which 24.4% resulted in death. By focusing on the exploration of explanatory factors for the occurrence of death during hospitalizations due to COVID-19, it contributes with relevant findings to the international debate, confirming knowledge that has been consolidated, raising questions and exposing specificities of the Brazilian context.

Sociodemographic factors and the presence of comorbidities have been identified as associated with COVID-19 hospitalization and death^20,21^. Similar to what was described in other studies, males, older age group gradually higher, black race/color, Charlson score, presence of Elixhauser comorbidity and obesity presented a higher adjusted odds of death^7, 8,22-24^.

Blacks were more likely to die during hospitalization in all estimated models, while browns showed significant positive differentiation only in models 1 and 2. A study that used the data from the Epidemiological Surveillance Information System (SIVEP-Gripe) to study hospital mortality related to COVID-19 in Brazil, between the end of February and the beginning of May, showed that being black or brown was the main risk factor for hospital mortality, with brown color presenting a higher risk of death than black^10^. These results are compatible with the Brazilian socioeconomic reality, where blacks and browns are more vulnerable to epidemics and other diseases because they live in worse health conditions, have less access to healthcare services, lower economic security levels, and, in the specific context of COVID-19, had to expose themselves more often to virus by going to the streets, using public transportation to get to workplaces and other crowded places in order to assure their livelihood^25^. Higher hospital mortality for blacks than whites has also been observed in international studies^26,27^. It is worth mentioning that some studies have been looking for explanations for the higher mortality among blacks due to pathophysiological mechanisms, and one of the relationships pointed out as deserving of future research are the relationships among being black, COVID-19 and greater risk of venous thrombosis^28^.

Hypertension and diabetes showed protective effect, which seems paradoxical and inconsistent with some reports in the literature^17,29^. However, these findings refer mainly to critical cases and the control by other variables may have modified their effect. On the other hand, in a review of the relationship between hypertension and the use of angiotensin-converting enzyme (ACE) inhibitors with COVID-19 outcomes, the authors argue that despite the potential role of immune responses in the pathogenesis of hypertension, there is no evidence to support the hypothesis that hypertension or inhibitors of the renin-angiotensin system contribute to unfavorable outcomes in viral infections. The study emphasizes that there is no current scientific basis for stating that hypertension or its treatment with drugs of the blocking class of the reninangiotensin system contribute to unfavorable results in COVID-19^30^. It is noteworthy that despite the low levels of comorbidity reporting in our data (only about 20% of hospitalizations), compared to what was reported by Petrilli et al. (2020), in which 80% of hospitalized patients had at least one of the chronic diseases examined, the strategies for measuring the comorbidities used were also associated with higher risk of death^20^.

Obesity has been identified as a risk factor. This finding is consistent with international studies that have shown that obesity is a risk factor for mortality among patients diagnosed with COVID-19, regardless of whether it is associated with other comorbidities and potential confounding factors^31^.

The median LOS was 5 days and the average was 6.9 days. These findings are consistent with a study conducted in the United States^17^, but differ from hospitalization data in Lombardy, the Italian region most affected by the pandemic in the first months of 2020, with a median of 28 days of hospitalization^32^. Differences in LOS may be associated with the age structure and degree of population aging in each location. In Brazil, the high risk of death in the first 24 hours of hospitalization stood out, which could be expressing the rapid level of health deterioration caused by the virus or problems in the journey to access health services in a timely manner or even the care that was immediately provided. Among the hospitalizations that used the ICU, the adjusted odds of death was extreme (OR 11.74), which seems to express predominantly the severity of the case, but also some synergy with the quality of care. In a meta-analysis that used 24 studies from three continents, a combined mortality of 41.6% of patients admitted to the ICU was observed, a value well below the 55.7% observed in this study. The study also observed that mortality in the ICU due to COVID-19 was higher than that normally observed in hospitalizations due to other viral pneumonias^4^.

In the context of the pandemic, the SUS hospital network has been crucial for responding to the demands for acute care that emerged. However, numerous problems related to supply and performance have arisen, especially the insufficient number of hospital beds and staff to perform specialized care in the ICU^8,33,34^, thus resulting in wide variation in effectiveness which is directly related to hospital mortality. The findings indicate significant differences between states and hospitals of different legal status classifications probably reflecting wide variations in the structure and general capacity of offering quality care. States located in the North region such as Acre, Roraima, Amazonas, Pará and Amapá presented, at the beginning of the COVID-19 outbreak, municipalities with exceptionally low or no capacity to treat severe cases of the disease^33^. This scenario is reflected in the comparison of the adjusted odds of death between patients hospitalized in Amapá (OR = 10.44), Acre (OR = 5.42) and Amazonas (OR = 3.07). In the Northeast, Rio Grande do Norte was the state with the highest odds of hospital death (OR = 2.64), counting with only three municipalities with minimal capacity to deal with severe cases of the infection at the beginning of the pandemic. Although the states of the South and Southeast regions present, on average, better supply of hospital beds, the worse outcomes observed in Rio de Janeiro (OR = 2.29) and São Paulo (OR = 1.24) compared to other states is worth mentioning. Both states were hard hit by the pandemic, illustrating the results’ variability and Rio de Janeiro was, in a certain period of the pandemic, the second in number of cases and recorded the highest fatality rates in the country. The high levels of case underreporting and low testing may be related to these findings^35^. The state also had serious problems related to management of the pandemic, poor planning of field hospitals as some were never completed and others were delivered too late.

Moreover, the variation in the chances of survival would be greater if the cases treated in the private system by those who have health insurance were counted here, where the higher supply and access to critical resources in specific states of the country generate unequal conditions for the prognosis of patients^36^. Beyond the inequalities, issues of political nature may have contributed to variations in performance between the states of São Paulo and Rio de Janeiro, particularly the articulation between the state and municipal government levels, and the recognition of the seriousness of the problem and responsibility towards citizens.

Our study has limitations. The main gap refers to the source of information used. SIH covers only the SUS hospital network, which makes it impossible to carry out a more comprehensive analysis including healthcare received by those privately insured. Additionally, the data flow from providers to the system, and its subsequent consolidation, is slower than desirable to monitor the care provided in a pandemic context that requires fast decisions. The sufficiency and quality of the information recorded should be stressed, in particular the high underreporting of comorbidities and the variable ‘race/color’. Further, it was not possible to include cases treated in the emergency wards, and data on the evolution of cases (such as vital signs), and on the care process (professionals involved, use of invasive mechanical ventilation and laboratory tests, including tests for the detection of COVID-19) are absent from this source, which impedes more specific analysis. The study also does not cover deaths that occurred outside hospitals, that may have also played an important role in understanding the real pandemic morbidity and mortality scenario.

It is important to remember that the pandemic has evolved dynamically throughout the country. This study addresses hospitalizations in the initial months. Faced with a scenario of cases’ spread in the interior of the country, it is likely that the same analyses in subsequent months provide another overview of how the states were affected.

Despite the limitations mentioned, the study has the merit of examining hospital mortality with national coverage, considering all COVID-19 patients who were admitted at hospitals and received care from SUS, capturing individual and contextual sociodemographic aspects and associated risk factors. Although the source of information, design and statistical modeling limits comparability, the findings corroborate those highlighted by Baqui et al. (2020) regarding regional and racial/ethnic variation in the Brazilian context^10^. In addition, although there is a vast and growing literature on COVID-19, there are still rare approaches with the strategies we used, focusing on the profile and outcomes of COVID-19 hospitalizations nationwide and exploring gradually the effects of groups of variables^10,17^. In the Brazilian context, the strong social gradient emerges, although information on race/color and geographic location is insufficient to trace the multiple facets of socioeconomic inequalities in the society^37^. The wide variability in the performance of the health system between states emerges. It can, in part, be attributed to the structure and prior organization of the services available, but it is also due to the insufficient regional/local capacity to coordinate actions to deal with COVID-19. The pandemic scenario in Brazil is also aggravated by the absence of national coordination at the federal level, which could be capable of mitigating the major regional differences in a continental and diverse country.

## Data Availability

The study is based on publicly accessible data.

## Acknowledgements

CCAP, MM e MCP are recipients of productivity fellowships from Conselho Nacional de Desenvolvimento Científico e Tecnológico (CNPq), Brazil

